# Men and loneliness in the Covid-19 pandemic: insights from an interview study with UK-based men

**DOI:** 10.1101/2021.06.30.21259536

**Authors:** John Ratcliffe, Paul Galdas, Mona Kanaan

## Abstract

As the SARS-COV-2 pandemic hit, the UK, like many countries, introduced severe restrictions on social contact, and injunctions of ‘social distancing’, to reduce transmission. This led to a concern that loneliness may increase, facilitating poorer mental and physical health. Twenty qualitative interviews were conducted, with a diverse group of UK-based men, between January and March 2021, during severe restrictions. Our aim was to generate new insights into men’s experience of loneliness during the pandemic, and consider the ramifications of these for continued/future restrictions, the easing of restrictions, and the future beyond the pandemic. Thematic analysis, focused on semantic themes, was employed as part of a ‘grounded’ epistemology whereby the stated perspectives of the interviewees drove the content of the study. Six themes were constructed: i) people to see and things to do (broken, changed, and new); ii) rethought and renewed recognition of what is important; iii) loneliness with a purpose; iv) loneliness as normal; v) anxiety of social contact; and vi) easier for some than others. The restrictions did cause some loneliness, particularly because of lost routines and opportunities, and the reduction in face-to-face interaction. However, a need to reduce transmission of SARS-COV-2, and a fear of catching it, became important features of participants lives that also affected loneliness and its causes. Remote forms of interaction were often utilised, and though they were imperfect, those that were dependable, were smaller than might be desired in person, and ensured a chance to speak, were constructed as more positive. The fear of Covid-19, and a loss of social skills, may mean that reducing restrictions alone will not return everyone to pre-pandemic levels of loneliness. Some believed the building of supportive local communities, and a destigmatisation of loneliness, may allow for the building of a more compassionate and less lonely society.

**What is known about this topic:**

- Loneliness is a public health concern, and is often a gendered experience.
- Pandemic related restrictions greatly reduced opportunities for social contact.
- We have a limited understanding of whether, and how, men experienced loneliness during the SARS-COV-2 pandemic.

**What this paper adds:**

- The loss of routines, and a lack of face-to-face interaction, were the greatest challenges to loneliness, particularly among solo-living men.
- Remote interaction may be better when routinised or dependable, in smaller groups, and with a structure facilitating the opportunity to speak.
- Anxiety of SARS-COV-2, and identities built in relation to reducing transmission, were important influences on the causes and severity of loneliness.

## 1 Introduction

Loneliness is commonly defined as a perceived lack, or loss, of meaningful social relationships (Townsend, 1957; Perlman and Peplau, 1981; Weiss, 1982; Cattan et al., 2005). In recent years, it has been extensively linked to poor mental and physical health (Cacioppo et al., 2006; Schinka et al., 2012; Bolton, 2012; Victor and Bowling, 2012; Valtorta et al., 2016a). Loneliness became a particular concern as ‘social distancing’ and ‘lockdowns’ became critical tools against SARS-COV-2 (Killgore et al., 2020; Brodeur et al., 2021).

In the months following the introduction of restrictions, some studies found a corresponding increase in loneliness (McQuaid et al. 2021; Bu et al. 2021), while others have not (Luchetti et al., 2020; Folk et al., 2020). Loneliness is a subjective emotion, often defined with reference to ‘social isolation’, which represents an objective lack of social contact (Perlman and Peplau, 1981; Cattan et al., 2005; Valtorta et al., 2016b). It is therefore theoretically plausible that ‘social distancing’ and ‘lockdown’ can increase ‘social isolation’ without increasing ‘loneliness’. Luchetti et al. (2020) found increased levels of social support, which they theorise may explain why restrictions did not lead to loneliness in their sample. However, little research has examined how and why this may be important in a pandemic, particularly for men, who may receive less emotional support than women (Stevens and Westerhof, 2006; McKenzie et al., 2018).

In China, Wang et al. (2020) found that, following the coronavirus outbreak, women were much more anxious than men, and in Slovenia, Kamin et al. (2020) found solo-living women were lonelier if there was a bigger gap between their pre-pandemic and post-pandemic amount of social contact. Numerous authors have suggested masculine identities eschewing outward signs of weakness facilitate an aggregate disinclination to acknowledge or seek help for loneliness (Nicolaisen and Thorsen, 2014; Yousaf et al. 2015; Rokach, 2018), yet whether and how this may be enacted in relation to a pandemic is unknown. For Franklin et al. (2019), ‘belongingness’, within gendered communities, is often key to preventing and alleviating loneliness. Though ‘social distancing’ may not be a sufficient condition for loneliness, it is likely to have affected men’s gendered agency and practices of ‘belongingness’. However, there is a paucity of research investigating men’s experiences. This study aims to investigate and highlight where and how men’s practices of belongingness, and experiences of loneliness, have changed for UK-based men, and the ramifications of this for policy, services, and wellbeing.

In England, a highly restrictive ‘lockdown’ was initiated in March 2020, in which people were ordered to ‘stay at home’ unless they were designated a ‘key worker’. This was gradually eased throughout the summer, and August saw the introduction of the ‘eat out to help out’ scheme, in which the government subsidised discounted meals in pubs and restaurants. However, increases in SARS-COV-2 cases through September (HM government 2021a) led to the introduction of a ‘tier’ system in October, which offered three different ‘levels’ of restrictions, but with the most restrictive tier less restrictive than the lockdown in March. In November, a four week nationwide ‘lockdown’ was enacted, albeit schools remained open, and the UK returned to the tier system in December. Initially, the government had created a five day ‘Christmas window’, in which people could visit and stay with families in their homes. This was then withdrawn, leading to many media sources suggesting the government had ‘cancelled Christmas’ (Murray, 2020). Shortly after Christmas, with hospitalisations and deaths still rising (HM Government 2021a), a nationwide lockdown legally similar to that of March 2020 was introduced once more.

The interviews that form the current study took place between January and March 2021, during this ‘lockdown’. The research questions were:

> *How has the pandemic affected men’s experiences of, and attitudes towards, loneliness?*
>
> *- what are the ramifications of their experiences and attitudes for easing restrictions, future pandemic situations, and a post pandemic world?*

## 2 Methods

Twenty semi-structured interviews, with men based across Northern England and Scotland, were conducted. Interviewees were sourced via an LGBTQ+ group, a sports centre, a community centre, a men’s activity group, an organisation promoting good health in black people, and organisations supporting voluntary work. The study employed a relatively ‘grounded’ approach, insofar as the content of this article was strongly driven by the interviewees (Charmaz, 1996). This paper focuses on data related to loneliness and the SARS-COV-2 pandemic (Braun and Clark 2006).

### 2.1 Sample

Participants were aged between 20 and 71, and were not required to have experienced loneliness. A maximum variation purposive sampling frame (Guest et al., 2013) was conducted to ensure some diversity. Table 1 lists demographic data. A ‘pragmatic’ approach was taken to interviewee numbers (Braun and Clark, 2021), as theoretical saturation was considered unfeasible (Low, 2019). Participants were contacted via a ‘gatekeeper’ in the organisations listed above, and gave written consent via email. A £10 gift voucher was offered for taking part. To maintain anonymity, demographic data is not linked to participant pseudonyms (Bell 2010; Ratcliffe et al. 2019).

**Table 1.** Demographic information of interviewees.

| Age | N (% of interviewees) |
| --- | --- |
| *18-30* | **5** (25) |
| *31-45* | **5** (25) |
| *46-60* | **7** (35) |
| *61+* | **3** (15) |
| **Ethnicity** |  |
| *White-British* | **14** (70) |
| *South-Asian* | **4** (20) |
| *Eastern-European* | **1** (5) |
| *White-African* | **1** (5) |
| **Sexual orientation** |  |
| *Heterosexual* | **12** (60) |
| *Bisexual* | **1** (5) |
| *Homosexual* | **7** (35) |
| **Gender orientation** |  |
| *Cisgender* | **19** (95) |
| *Transgender* | **1** (5) |
| **Attended higher education** |  |
| *Yes, in the UK* | **5** (25) |
| *Yes, in another country* | **2** (10) |
| *Current student* | **3** (15) |
| *No* | **10** (50) |
| **Living situation** |  |
| *Solo-living* | **8** (40) |
| *With spouse/partner (with or without children)* | **7** (35) |
| *With parents/guardians* | **4** (20) |
| *With housemates* | **1** (5) |

### 2.2 Data collection

Interviews were conducted remotely, by John Ratcliffe, via video call (Google hangouts, Zoom) or telephone, in an enclosed room using a headset. They lasted between 30 and 120 minutes, and were video-recorded and auto-transcribed, or recorded on the telephone and manually transcribed. The interviews loosely consisted of three parts. First, a less structured interview, broadly discussing loneliness, was employed. This aimed to utilise Hollway and Jefferson’s (2000; 2008) technique of ‘free association’, a method aiming to allow participants to frame topics according to their own discursive associations, therefore is congruent with a ‘grounded’ epistemology. Secondly, to ensure focused reflexivity on the impact of the pandemic, participants were asked whether and how the pandemic had affected them (Mason, 2002). Finally, questions specifically related to maleness, masculinities, and loneliness were pursued, again to ensure a reflexive account.

### 2.3 Analysis

Thematic analysis, focused on specific data relevant to the pandemic and loneliness, was employed (Braun and Clark, 2006). The results focused on constructing semantic ‘surface’ level themes (Braun and Clark, 2006). This helped to ensure the results had a clear ‘link’ to the data (Vindrola-Padros and Johnson, 2020), assisting the employment of a ‘grounded’ epistemology. It also allowed the study to be conducted rapidly, without sacrificing rigour, allowing for a timelier addition to pandemic literature (Vindrola-Padros et al. 2020). Open coding was employed, then built and narrowed into specific and consistent themes (Moghaddam, 2006). Coding was conducted in NVivo (2020), in six stages:

1. Interviews were digitally recorded, transcribed, and uploaded to Nvivo.
2. Open coding was conducted, whereby data was assigned a large number of descriptive labels broadly related to loneliness. The decision to produce an analysis focused on the pandemic was taken after this stage.
3. A second open coding was conducted, which built a large number of new codes solely related to the pandemic and loneliness.
4. Codes were reviewed, adapted, and narrowed.
5. Themes were built, defined, and named.
6. Themes were adapted and reproduced into an article suitable format.

## 3 Results

Six themes were constructed. ‘People to see and things to do: broken, changed, and new’ (3.1) summarises how the men’s lives and social connections were disrupted and reformed, particularly around more ‘covid-safe’ activities and in narrowed social spheres focused on the home and local community. ‘Rethought and renewed recognition of what is important’ (3.2) exemplifies how these disruptions led many to reconsider what about their lives and social connections is important, particularly when contrasting ‘face-to-face’ with remote forms of social interaction. ‘Loneliness with a purpose’ (3.3) emphasises the moral imperative to stop transmission of SARS-COV-2, and how that impacted their emotional experiences. ‘Loneliness as normal’ (3.4) represented a perception that loneliness was to be expected during the restrictions. ‘Anxiety of social contact’ (3.5) consisted of a fear of catching the virus, and of an anxiety wrought by limited social contact. Lastly, ‘easier for some than others’ (3.6) aimed to capture how the men often discussed groups for whom the situation is more difficult. Despite this, the themes listed were relatively universal, and only tended to differ in terms of magnitude and method. For example, where some men’s lives were greatly changed, others relayed it changed a little, and where some rarely attended group video calls, others did so regularly. It was at this level the pandemic became ‘easier for some than others’. Most notably, some solo-living men by the loss of wider social spheres, poorer men appeared less likely to experience a positive sense of ‘local community’, and men who are more at risk of Covid-19 were particularly anxious about social contact.

### 3.1 People to see and things to do: broken, changed, and new

> *Being a creature of habit, there are things that repeat…and this last year has just broken that cycle (DA)*.

The men often emphasised the disruption to their routines and usual social opportunities. Keeping ‘busy’ was often stated as critical to preventing loneliness, and the pandemic had greatly intervened in this. Many humorously emphasised home-centred activities such as DIY and cleaning as ways of staving off boredom:

> *Some of the humour shared has been relating to…the number of times that somebody else has cleaned the bathroom with an intensity that they’d never have done before. So I think there’s, at least in a sort of trivial way, using humour, there’s an understanding and discussion of loneliness (DJ)*.

Though constructed as banal, these activities could distract, and, more importantly, could form a narrative of solidarity and support.

Many sought new activities and routines, and, unlike those above, these were often constructed as important, particularly when they were routinised or dependable. DA began a daily running routine through a phone app, and described this as having ‘saved’ him from loneliness. Remote social contact was frequently stated to be a relatively poor substitute for what was often termed ‘face to face’ socialising (JM, HJ, VP, SB, GT, SM), yet most emphasised it as worth engaging with. SB, for example, spoke to his brother on the telephone every evening, and GT spoke highly of an LGBTQ+ support group. Remote interaction also provided some people with new opportunities. SC, who had a limiting physical disability, noted that ‘I probably have more contact with people now that I did before’, and was even invited to speak at an event held in the USA.

Many emphasised that their social spheres had narrowed into a focus on home environments and local communities. For some solo-living participants, particularly gay men who, in this study, were more likely to have built their social connections in public spheres, this was difficult:

> *People complaining about…a partner, or the kids are driving me up the wall and stuff, and I think, well, swap with me for a week, see what it is (NT)!*

This was also a problem for those who were seeking sexual and/or romantic partners, as opportunities to meet people were greatly reduced, and younger participants who lived with family, such that one called it a ‘pressure cooker environment’ (OS). However, this narrowing of spheres could also facilitate stronger relationships with existing partners, family, and friends, and several men expressed a deep gratitude for this. Furthermore, some spoke with enthusiasm for an improved ‘local community’. This appeared to be parallel to narrowed spheres insofar as it was rooted in specific localities, particularly voluntary groups, and neighbours, yet also seemed to represent a more abstract sense of social support and connection. Furthermore, there were concerns this was fading, or temporary - ‘how many people will carry on that work, and how much will stop’ (SC)?

### 3.2 Rethought and renewed recognition of what is important

> *This whole period has been really cathartic because it’s allowed me to figure out what it is that does make me happy. Figure out what’s good about me, figure out that I am worth enough on my own, I don’t need to have somebody else to validate me (NT)*.

Many of the men expressed a renewed gratefulness and/or appreciation of relatively good aspects of their life, such as good health, having close relationships, economic comfort, and outdoor spaces, particularly green spaces. However, restrictions also intervened with actualising newfound perspectives on what is important. DJ, for example, had realised the importance of being ‘somewhere else’, instead of at home, yet his opportunities to do so were limited.

Many of the men discussed a newfound awareness of what constituted a positive social interaction. This was usually constructed by contrasting ‘face-to-face’ interaction with remote forms of interaction, particularly video-calls. In remote interaction, participants relayed a lack of physical intimacy, difficulties with understanding context through body language, greater anxiety, and difficulties with being able to get involved in conversations (often because one individual would dominate). Nevertheless, several relayed how remote interactions could improve, suggesting smaller groups, adequate opportunity to take part, and dependable availability. Overall, it seemed dependable socialising, in which they felt comfortable and able to take part, was key to ‘good’ interaction, and that this was easier with ‘face-to-face’ interaction.

### 3.3 Loneliness with a purpose

Some emphasised that they understood the rationale of the restrictions, and that it had a bearing on their emotional experiences:

> *There’s a friend of mine who’s on the covid ward…and you think what they’re going through compared to what I’m doing, basically just sitting doing nothing, I can deal with that. Couldn’t deal with what he does, but my tiny little bit of help, just to do nothing really, it’s not that much to ask (SB)*.

VP described this as loneliness with a ‘purpose’, and noted there was a positive aspect to this as it gave meaning to his life. As a result, he felt particularly lonely after being invited to attend a Christmas party:

> *…if I get an invitation, which I got several times, I have to say like bloody hell don’t you read a newspaper?! There’s another lockdown! And it makes me feel sorry to explain to you it’s inappropriate (VP)*.

For some participants, then, being physically alone represented an act of social benefit, therefore loneliness was less of a problem. When others failed to share this social cause, though, some felt lonely, even if, as in VP’s statement above, they were being invited to spend time with other people.

### 3.5 Loneliness as normal

Several participants expressed concerns that the restrictions might lead to greater loneliness across society. Nevertheless, some also hoped that this may help to destigmatise loneliness, creating a better future. Notably, men who had not experienced increased loneliness appeared to consider it surprising, even shameful:

> *Interestingly, the covid thing hasn’t really affected me (BR)*.
>
> *It’s not been that much different (HY)*.
>
> *I feel a bit guilty about admitting that I’m alright (VP)*.

During pandemic restrictions, then, loneliness was often constructed as an expected state, distinct from a rarer and more highly stigmatised state prior to the pandemic.

### 3.5 Anxiety of social contact

Two sub-themes in people’s anxieties of social contact were identified. The first consisted of a fear of catching SARS-COV-2, and was particularly salient in higher risk participants. HJ, who had a compromised immune system, stated he was ‘paranoid about going near people’, and later mentioned that sitting in a restaurant would leave him ‘uncomfortable and vulnerable’. In this way, HJ was ‘choosing’ to avoid social contact, regardless of restrictions, yet it was a choice heavily influenced by the severity of the risk to his health, and still represented a lonely experience. The second sub-theme was a newfound broader anxiety. SC, for example, noted he and others might ‘get into a routine and forget about all the people’, and NT worried that he would lose his social ‘skills’ and find it hard to ‘integrate back into society’.

### 3.6 Easier for some than others

As noted, some solo-living men framed the situation as particularly difficult, yet others, such as BR, VP, and HY, did not. Some older participants ruefully mentioned difficulties with, and dislikes of, remote interactions, and related this to their age. More commonly, though, the men in this study believed they found the situation *easier* than others did. Some believed the restrictions may be more difficult for younger people, particularly children, as it is a time where people ‘develop within a social group’ (VP). However, the younger participants in this study did not identify this. Most surprisingly, perhaps, some participants with past experiences of loneliness, or other mental health problems, believed this an advantage, as it prepared them for the situation:

> *People who suddenly couldn’t have what they always had couldn’t get their heads around why they couldn’t have it anymore. But I was already on that journey before because I lost all of that before I got into my recovery (JM)*.

HZ, a Pakistani man living in what he described as a ‘deprived’ area, made numerous observations that appeared consistent with other interviews. Firstly, he believed older BME people tend to receive more attention from their children, which reduced their loneliness during the pandemic. Notably, South Asian interviewees in this study all spoke of regular and intimate social contact with children and/or parents, often because they lived in fluid multi-generational housing. However, HZ also believed BME people tended to be less trusting of services, thus may be less likely to receive pandemic-related assistance. This did not feature specifically in other interviews, although CO was critical of support services, particularly care homes. Lastly, HZ noted that poorer people often live in less ‘positive’ local communities, where crime and low self-worth are more common. Where some men emphasised positive ‘local communities’ during the pandemic (JM, SC), then, it may be important that others, such as GT and HY, framed their community in relation to drugs, violence, and distrust.

## 4 Discussion

Little is known about men’s experiences of loneliness during the Covid-19 pandemic. This study provides new insights derived from qualitative interviews with men during a period of severe restrictions. Restrictions sometimes facilitated loneliness, but this only told part of the story. As in previous pandemics, SARS-COV-2 appeared to have destabilised people’s social structures (Strong, 1990; Cava et al., 2005), and interrupted routines and opportunities were a key cause of loneliness. However, SARS-COV-2 has lasted longer than the periods addressed by Strong and Cava et al, perhaps explaining why this study found more signs of new routines and behaviours. Indeed, narrowed social spheres may result in smaller, but closer, social networks for years, particularly if people are more anxious of social settings and/or have lost social skills.

During times where restrictions are being eased, people may need to balance anxieties, and new routines, against their preference for ‘face-to-face’ interaction, and possibly against a ‘fear of missing out’ (Baker et al., 2016). This may be particularly pertinent for those who have constructed their isolation as ‘loneliness with a purpose’, given that it emphasises restricting in-person contact as a communal moral imperative. Time, vaccines, and lower case rates may change these psychologies.

Nevertheless, community services of all kinds may need to move gradually, take covid-cautious approaches, and communicate with people in a manner acknowledging these anxieties. It also suggests an emphasis on ‘personal responsibility’ (Williams 2021) may be difficult for some, given a complex backdrop of anxiety, lost social skills, and moral imperatives reformed in narrowed social spheres.

Williams et al. (2020) reviewed covid-friendly interventions to alleviate loneliness, and found the strongest evidence for ‘therapeutic’ interventions such as mindfulness, as well as some evidence that maintaining and reconnecting with existing social networks may be more effective than facilitating new social relationships. In this study, the renewed sense of what is ‘important’, and the idea that previous mental health struggles facilitated better resilience, offer further evidence for ‘therapeutic’ approaches, and the comfort derived from spouses, partners, family, and close friends mirrors their suggestion that existing networks may be more beneficial than new ones. The study also offers support for allowing outdoor exercise, and ‘support bubbles’ for solo-living people (HM Government 2021b), during severe restrictions, given that outdoor exercise and being ‘somewhere else’ were framed as important for alleviating loneliness, and solo-living could be particularly challenging.

Noone et al. (2020) reviewed the efficacy of services using remote forms of interaction to alleviate loneliness in older people, and found mixed results. In this study, participants discussed numerous problems and disadvantages to remote interaction, yet also suggested a number of ways it can be good, perhaps explaining the heterogeneity in Noone’s review. Specifically, it seemed that smaller groups, where people felt involved and able to speak, and were dependable in terms of their availability, were most beneficial. However, they remained a less positive substitute for ‘face-to-face’ interaction, therefore long term use without face-to-face interactions may constitute a component of ‘lockdown fatigue’ (Mahase 2020; Goldstein et al. 2021).

Ratcliffe et al. (2019) posit that older men may place a sense of ‘social worth’ as critical to preventing loneliness, and emphasise that this does not always require social contact. The notion of ‘loneliness with a purpose’ may represent a stark example of this, although Kamin et al.’s (2021) study of solo-living women in Slovenia also found they related a moral responsibility to reduce transmission, albeit without suggesting this reduced loneliness. In contradiction to this study, research from Mind (2020) and Gillard et al. (2021) found that BME people, and those with mental health problems, faced additional psychological difficulties. Bartholomaeus and Tarrant (2016) suggest that older men may construct a masculine identity as a ‘sage’, therefore the notion that past experiences of loneliness make it easier to cope with the pandemic may represent a masculine identity as a ‘sage’. Nevertheless, it appeared to build genuine resilience for some of the men in this study.

Numerous studies have suggested men are worse affected by not having a spouse/partner than women (Pinquart and Sorensen, 2001; Bergland et al. 2016; Nowland et al., 2018). This may explain why some solo-living men, who previously had more public facing lives, were particularly negatively affected. However, Kamin et al. (2021) found a similar difficulty among women who had more public facing lives, thus rather than being gendered, this may parallel Folk et al.’s (2020) finding that ‘extroverts’ are more affected than ‘introverts’. Indeed, if men were aggregately more reliant on spouses/partners prior to the pandemic, and social spheres have narrowed towards the home, this may explain why some research has suggested women have been more negatively affected (Wang et al., 2020). This may also why solo-living men identifying as gay were more affected than heterosexual solo-living men - homosexuality has been placed as inherently incompatible with patriarchal goals of a heterosexual nuclear family (Connell, 2005), hence a greater tendency towards social spheres outside of the home. Furthermore, if ‘loneliness with a purpose’ reduces negative emotional affects in men more than it does in women, this could also explain worse affects in women. Finally, if loneliness is ‘normal’ during covid restrictions, it could reduce the likeliness of men understating loneliness (Nicolaisen and Thorsen, 2014; Yousaf et al. 2015; Rokach, 2018), although seeking help for it may still represent a vulnerable position to be avoided (Yousaf et al. 2015). These gendered interpretations of the results, though, are highly theoretical, and require further research.

### 4.1 Strengths and limitations

This study cannot gauge the scale of these themes across societies (Bryman, 2006), and it is difficult to identify precisely whether and how these findings are gendered in this data. The participants were fairly diverse, yet none were black, single parents, or either under 20 or over 71 years old. Though participants suggested numerous ways the pandemic was ‘easier for some than others’, these were often vague assertions, usually related to groups other than themselves, therefore this requires more focused research. The focus on semantic themes may also limit insight (Braun and Clarke, 2006). No participant had experienced Covid-19, and only one participant mentioned a person they knew who had. As such, the study offers limited insight to people with lived experience of the virus, particularly bereaved people who may be at risk of loneliness (Stroebe and Schut, 2020). The study is unable to balance people’s needs in relation to loneliness against the need to avoid transmission of SARS-COV-2. Support bubbles, outdoor exercise, attention from (adult) children, and involvement in local communities, were constructed as helpful to loneliness, yet involve social contact that may increase transmission. Conversely, it is impossible to derive from this data whether, when, and to what extent, anxiety of SARS-COV-2 is rational emotional response, or a cognitive problem.

By focusing on semantic themes, this study was able to provide a timely addition to the literature, with a grounded focus on people’s stated experiences during a pandemic at its height. Though restrictions could cause loneliness, conceptualising the problem as ‘restrictions = loneliness’ was insufficient. Rather, *the pandemic significantly disrupted people’s agency in affecting loneliness, and facilitated changes to their discursive practices of what causes and constitutes it*. During restrictions, social opportunities and routines were broken, and the loss of face-to-face interactions was particularly salient. Nevertheless, new social opportunities and routines, a sense of local community, and a clear understanding of the ‘purpose’ of the restrictions, that was understood and respected by others, could do much to alleviate loneliness. Remote forms of communication were imperfect, but they could be positive, and held significant capacity for improvement. Though easing restrictions has obvious potential for reintroducing valued ‘face-to-face’ contact, it may be important to be aware of people’s anxieties, newfound discursive practices, and lost social skills. Lastly, several notes of hope were constructed, particularly around a ‘normalisation’ of loneliness and local communities, which some felt may aid in building a more open, fair, and compassionate society.

## Data Availability

The data are not available due to privacy or ethical restrictions.

## References

Baker, Z. G., Krieger, H., & LeRoy, A. S. (2016). Fear of missing out: Relationships with depression, mindfulness, and physical symptoms. Translational Issues in Psychological Science, 2(3), 275.

Bell, J. (2010). Doing your Research Project: A guide for first time researchers in education, health and social science (5thed.). Maidenhead: Open University Press.

Bergland, A. M. G., Tveit, B., & Gonzalez, M. T. (2016). Experiences of older men living alone: A qualitative study. Issues in mental health nursing, 37(2), 113–120.

Bolton, M. (2012). Loneliness–the state we’re in. A Report of Evidence Compiled for the Campaign to End Loneliness. Oxfordshire: Age UK, Oxfordshire.

Braun, V., & Clarke, V. (2006). Using thematic analysis in psychology. Qualitative research in psychology, 3(2), 77–101.

Braun, V., & Clarke, V. (2021). To saturate or not to saturate? Questioning data saturation as a useful concept for thematic analysis and sample-size rationales. Qualitative research in sport, exercise and health, 13(2), 201–216.

Brodeur, A., Clark, A. E., Fleche, S., & Powdthavee, N. (2021). COVID-19, lockdowns and well-being: Evidence from Google Trends. Journal of public economics, 193, 104346.

Bryman, A. (2016). Social research methods. Oxford university press.

Bu, F., Steptoe, A., & Fancourt, D. (2020). Loneliness during a strict lockdown: Trajectories and predictors during the COVID-19 pandemic in 38,217 United Kingdom adults. Social Science & Medicine, 265, 113521.

Cacioppo, J. T., Hughes, M. E., Waite, L. J., Hawkley, L. C., & Thisted, R. A. (2006). Loneliness as a specific risk factor for depressive symptoms: cross-sectional and longitudinal analyses. Psychology and aging, 21(1), 140.

Cattan, M., White, M., Bond, J., & Learmouth, A. (2005). Preventing social isolation and loneliness among older people: a systematic review of health promotion interventions. Ageing & Society, 25(1), 41–67.

Cava, M. A., Fay, K. E., Beanlands, H. J., McCay, E. A., & Wignall, R. (2005). The experience of quarantine for individuals affected by SARS in Toronto. Public Health Nursing, 22(5), 398–406.

Charmaz, K. (2006). Constructing grounded theory: A practical guide through qualitative analysis. London: Sage.

Connell, R. W. (2005). Masculinities. Polity.

Folk, D., Okabe-Miyamoto, K., Dunn, E., Lyubomirsky, S., & Donnellan, B. (2020). Did social connection decline during the first wave of COVID-19?: the role of extraversion. Collabra: Psychology, 6(1).

Franklin, A., Barbosa Neves, B., Hookway, N., Patulny, R., Tranter, B., & Jaworski, K. (2019). Towards an understanding of loneliness among Australian men: Gender cultures, embodied expression and the social bases of belonging. Journal of sociology, 55(1), 124–143.

Goldstein, P., Yeyati, E., & Sartorio, L. (2021). Lockdown fatigue: The diminishing effects of quarantines on the spread of COVID-19. Covid Economics, 67, 1–23.

Guest, G., Namey, E. E., & Mitchell, M. L. (2013). Collecting qualitative data: A field manual for applied research. Sage.

HM Government (2021a) UK Summary. https://coronavirus.data.gov.uk

HM Government (2021b) Making a support bubble with another household. https://www.gov.uk/guidance/making-a-support-bubble-with-another-household

Hollway, W., & Jefferson, T. (2000). Doing qualitative research differently: Free association, narrative and the interview method. London: Sage.

Hollway, W., & Jefferson, T. (2008). The free association narrative interview method. In: Given, LM. ed. The SAGE Encyclopedia of Qualitative Research Methods. Sevenoaks, California: Sage, pp. 296–315.

Kamin, T., Perger, N., Debevec, L., & Tivadar, B. (2021). Alone in a Time of Pandemic: Solo-Living Women Coping With Physical Isolation. Qualitative Health Research, 31(2), 203–217.

Killgore, W. D., Cloonan, S. A., Taylor, E. C., & Dailey, N. S. (2020). Loneliness: A signature mental health concern in the era of COVID-19. Psychiatry research, 290, 113117.

Low, J. (2019). A pragmatic definition of the concept of theoretical saturation. Sociological Focus, 52(2), 131–139.

Luchetti, M., Lee, J. H., Aschwanden, D., Sesker, A., Strickhouser, J. E., Terracciano, A., & Sutin, A. R. (2020). The trajectory of loneliness in response to COVID-19. American Psychologist.

Mahase, E. (2020). Covid-19: Was the decision to delay the UK’s lockdown over fears of “behavioural fatigue” based on evidence?. BMJ. 370. doi: https://doi.org/10.1136/bmj.m3166

McKenzie, S. K., Collings, S., Jenkin, G., & River, J. (2018). Masculinity, social connectedness, and mental health: Men’s diverse patterns of practice. American journal of men’s health, 12(5), 1247–1261.

McQuaid, R. J., Cox, S. M., Ogunlana, A., & Jaworska, N. (2021). The burden of loneliness: implications of the social determinants of health during COVID-19. Psychiatry research, 296, 113648.

MIND (2020) Existing inequalities have made mental health of BAME groups worse during pandemic, says Mind. https://www.mind.org.uk

Moghaddam, A. (2006). Coding issues in grounded theory. Issues in educational research, 16(1), 52–66.

Murray, W. (2020) ‘Christmas cancelled’: what the papers say as UK Covid bubbles burst. The Guardian. https://www.theguardian.com

Nicolaisen, M., & Thorsen, K. (2014). Who are lonely? Loneliness in different age groups (18–81 years old), using two measures of loneliness. The International Journal of Aging and Human Development, 78(3), 229–257.

Noone, C., McSharry, J., Smalle, M., Burns, A., Dwan, K., Devane, D., & Morrissey, E. C. (2020). Video calls for reducing social isolation and loneliness in older people: a rapid review. Cochrane Database of Systematic Reviews, (5).

Nowland, R., Talbot, R., & Qualter, P. (2018). Influence of loneliness and rejection sensitivity on threat sensitivity in romantic relationships in young and middle-aged adults. Personality and Individual Differences, 131, 185–190.

NVivo (2020). QSR International Pty Ltd. https://www.qsrinternational.com/nvivo-qualitative-data-analysis-software/home

Perlman, D., & Peplau, L. A. (1981). Toward a social psychology of loneliness. Personal relationships, 3, 31–56.

Pinquart, M., & Sorensen, S. (2001). Influences on loneliness in older adults: A meta-analysis. Basic and applied social psychology, 23(4), 245–266.

Ratcliffe, J., Wigfield, A., & Alden, S. (2019). ‘A lonely old man’: empirical investigations of older men and loneliness, and the ramifications for policy and practice. Ageing & Society, 1–21.

Rokach, A. (2018). The effect of gender and culture on loneliness: A mini review. Emerging Science Journal, 2(2), 59–64.

Schinka, K. C., VanDulmen, M. H., Bossarte, R., & Swahn, M. (2012). Association between loneliness and suicidality during middle childhood and adolescence: longitudinal effects and the role of demographic characteristics. The Journal of psychology, 146(1-2), 105–118.

Stevens, N., & Westerhof, G. J. (2006). Marriage, social integration, and loneliness in the second half of life: A comparison of Dutch and German men and women. Research on Aging, 28(6), 713–729.

Strong, P. (1990). Epidemic psychology: a model. Sociology of Health & Illness, 12(3), 249–259.

Townsend, P. (1957). The family life of old people; an inquiry in East London. London: Routledge & K. Paul.

Valtorta, N. K., Kanaan, M., Gilbody, S., & Hanratty, B. (2016b). Loneliness, social isolation and social relationships: what are we measuring? A novel framework for classifying and comparing tools. BMJ open, 6(4), e010799.

Valtorta, N. K., Kanaan, M., Gilbody, S., Ronzi, S., & Hanratty, B. (2016a). Loneliness and social isolation as risk factors for coronary heart disease and stroke: systematic review and meta-analysis of longitudinal observational studies. Heart, 102(13), 1009–1016.

Victor, C. R., & Bowling, A. (2012). A longitudinal analysis of loneliness among older people in Great Britain. The Journal of psychology, 146(3), 313–331.

Vindrola-Padros, C., Chisnall, G., Cooper, S., Dowrick, A., Djellouli, N., Symmons, S. M., … & Johnson, G. A. (2020). Carrying out rapid qualitative research during a pandemic: emerging lessons from COVID-19. Qualitative health research, 30(14), 2192–2204.

Vindrola-Padros, C., & Johnson, G. A. (2020). Rapid techniques in qualitative research: A critical review of the literature. Qualitative Health Research, 30(10), 1596–1604.

Wang, Y., Di, Y., Ye, J., & Wei, W. (2021). Study on the public psychological states and its related factors during the outbreak of coronavirus disease 2019 (COVID-19) in some regions of China. Psychology, health & medicine, 26(1), 13–22.

Weiss, R. S. (1982). Attachment in adult life. The place of attachment in human behavior, 171–184.

Williams, S. (2021) The UK’s coronavirus policy still places too much responsibility—and blame—on the public. BMJ. doi: https://doi.org/10.1136/bmj.n1373

Williams, C. Y., Townson, A. T., Kapur, M., Ferreira, A. F., Nunn, R., Galante, J., … & Usher-Smith, J. A. (2021). Interventions to reduce social isolation and loneliness during COVID-19 physical distancing measures: A rapid systematic review. PloS one, 16(2), e0247139.

Yousaf, O., Popat, A., & Hunter, M. S. (2015). An investigation of masculinity attitudes, gender, and attitudes toward psychological help-seeking. Psychology of Men & Masculinity, 16(2), 234.

